# How closely is COVID-19 related to HCoV, SARS, and MERS? : Clinical comparison of coronavirus infections and identification of risk factors influencing the COVID-19 severity using common data model (CDM)

**DOI:** 10.1101/2020.11.23.20237487

**Authors:** Yeon Hee Kim, Ye-Hee Ko, Sooyoung Kim, Kwangsoo Kim

## Abstract

**Background:** South Korea was one of the epicenters for both the 2015 Middle East Respiratory Syndrome and 2019 COVID-19 outbreaks. However, there has been a lack of published literature, especially using the Electronic Medical Records (EMR), that provides a comparative summary of the prognostic factors present in the coronavirus-derived diseases. Therefore, in this study, we aimed to evaluate the distinct clinical traits between the infected patients of different coronaviruses to observe the extent of resemblance within the clinical features and to identify unique factors by disease severity that may influence the prognosis of COVID-19 patients.

**Methods:** We utilized the common data model (CDM), which is the database that houses the standardized EMR. We set COVID-19 as a reference group in comparative analyses. For statistical methods, we used Levene’s test, one-way Anova test, Scheffe post-hoc test, Games-howell post-hoc test, and Student’s t-test for continuous variables, and chi-squared test and Fisher’s exact test for categorical variables. With the variables that reflected similarity in more than two comparisons between the disease groups yet significantly different between the COVID-19 severity groups, we performed univariate logistic regression to identify which common manifestations in coronaviruses are risk factors for severe COVID-19 outcomes.

**Findings:** We collected the records of 2840 COVID-19 patients, 67 MERS patients (several suspected cases included), 43 SARS suspected patients, and 87 HCoV patients. We found that a significantly higher number of COVID-19 patients had been diagnosed with comorbidities compared to the MERS and HCoV groups (48.5% vs. 10.4 %, p < 0.001 and 48.5% vs. 35.6%, p < 0.05) and also that the non-mild COVID-19 patients reported more comorbidities than the mild group (55.7% vs. 47.8%, p < 0.05). There were overall increases in the levels of fibrinogen in both sets of disease and severity groups. The univariate logistic regression showed that the male sex (OR: 1.66; CI: 1.29-2.13, p < 0.001), blood type A (OR: 1.80; CI: 1.40-2.31, p < 0.001), renal disease (OR: 3.27; CI: 2.34-4.55, p < 0.001), decreased creatinine level (OR: 2.05; CI: 1.45-2.88, p < 0.001), and elevated fibrinogen level (OR: 1.59, CI: 1.21-2.09, p < 0.001) are associated with the severe COVID-19 prognosis, whereas the patients reporting gastrointestinal symptoms (OR: 0.42; CI: 0.23-0.72, p < 0.01) and increased alkaline phosphatase (OR: 0.73; CI: 0.56-0.94, p < 0.05) are more less likely to experience complications and other severe outcomes from the SARS-CoV-2 infection.

**Interpretation:** The present study observed the highest resemblance between the COVID-19 and SARS groups as clinical manifestations that were present in SARS group were linked to the severity of COVID-19. In particular, male individuals with blood type A and previous diagnosis of kidney failure were shown to be more susceptible to developing the poorer outcomes during COVID-19 infection, with a presentation of elevated level of fibrinogen.

## Background

COVID-19, a global pandemic that has caused more than 2 million deaths and over 90 million cases since its outbreak^1^, is a novel respiratory viral disease caused by the SARS-CoV-2 virus that was first discovered in Wuhan, China, in December of 2019^2-5^. Coronaviruses are the RNA viruses that are commonly present in bats and belong to the family *Coronaviridae*. The family comprises four genera, *Alphacoronavirus, Betacoronavirus, Gammacoronavirus*, and *Deltacoronavirus*, of which the two, *Alphacoronavirus* and *Betacoronavirus* are known to cause respiratory infections in humans. In addition to the COVID-19, there are six other human coronaviruses that have been previously reported since the 1960s^6^, including HCoV229E, HCoV-NL63, HCoV-OC43, HCoV-HKU1, SARS-CoV-1, and MERS-CoV. While the HCoV strains cause mild upper respiratory diseases, the zoonotic strains including the SARS-CoV-1, MERS-CoV, and SARS-CoV-2 cause severe respiratory symptoms and complications with high mortality rates^6-15^. With the continuum of emergency outbreaks from coronavirus, a comparative assessment of the clinical traits between the infections and a stratification of common clinical variables by severity of the novel COVID-19 can be, therefore, useful in determining the specific at-risk population of each disease, risk factors of infection and severity, and degree of resemblance between the coronaviruses, in order to establish a comprehensive set of data. Thus, in this study, we aim to perform, using the real-world data of Seoul National University Hospital, a clinical characterization of the infected and suspected South Korean patients of HCoV strains, SARS-CoV, MERS-CoV, and SARS-CoV-2, and observe whether any common or distinct trait observed between the diseases may significantly influence the prognosis of the COVID-19 patients.

## Methods

### I. Data source

We designed a retrospective cohort observational study using Observational Medical Outcomes Partnership Common Data Model (OMOP CDM) based in Seoul National University Hospital located in Seoul, South Korea. CDM is the database that houses the Electronic Medical Records (EMR) transformed into the common format to be used by the multiple institutions for research purposes. It is the global data network shared by 19 different countries with more than half a billion patient records altogether^16^. The platform provides deidentified patient records of demographics, diagnosis, hospital admission, provider, prescription, and laboratory values, and vital statues^16^. Due to the privacy policy, there was no patient mortality information in our database. There was no patient or public involvement in this study. The study was approved by the Seoul National University Hospital Institutional Review Board (Seoul, South Korea) on April 2^nd^, 2020.

### II. Study Cohort

The follow-up period was from October 15^th^ of 2004 to July 31^st^ of 2020. Without any restriction on age and gender, we collected the records of symptoms, comorbidities, and laboratory test results of patients who were diagnosed with HCoV229E, HCoV-NL63, HCoV-OC43, HCoV-HKU1 (HCoVs), SARS-CoV-1, MERS-CoV, and SARS-CoV-2 infections using International Classification of Diseases, Tenth Revision (ICD-10)^17^. The index date of virus infection was determined by the first visit to the hospital. It is to note that the PCR results were not available, and the study cohorts were established solely on the diagnoses from the EMR. Therefore, SARS-CoV-1 group in this study involves the suspected individuals who showed clinical symptoms of SARS at the time of their hospital visits and had been diagnosed as SARS. MERS-CoV group also includes the several suspected cases. We divided the COVID-19 patients by their disease severity based on the published criteria from the World Health Organization (WHO)^18^. Among the COVID-19 patients, those who experienced mild cold-like symptoms with no existing pneumonia were classified as mild, whereas the patients who reported any clinical presentation of pneumonia only or with oxygen saturation (SpO_2_) under 90% were classified as non-mild. Laboratory test results were divided into low, normal, and high levels with a reference to published ranges by Seoul Clinical Laboratories^19^ (Appendix 1).

### III. Statistical analysis

There are total three statistical analyses in this study. First, we conducted a comparative clinical characterization of four disease groups with COVID-19 group as a reference. Continuous variables were expressed by the mean values with ± standard deviations (SD) with the appropriate units whereas categorical variables were represented by frequency and percentage (%). The Levene’s test, one-way Anova test, Scheffe post-hoc test, Games-howell post-hoc test, and Student’s t-test were used to compare continuous variables, and the χ^2^ test or Fisher’s exact test were used for the categorical variables^20-25^. With the same set of variables, listed analyses above were performed to compare between the mild and non-mild COVID-19 groups. To perform a univariate logistic regression^26-27^ to explore the extent of influence by risk factors on severity of COVID-19, we set two criteria for a variable selection. First, we considered statistical similarity between the variables of disease groups to reflect clinical resemblance between the patients. Second, among the variables that showed such similarity, we selected the ones with p-values less than 0.05 in severity analyses. The regression model was adjusted for age and gender. We excluded any variable that had null or sparse patient records. All statistical analyses were carried using R version 3.6.2.

## Results

### I. Comparative analyses between the coronavirus infections

#### Demographic and clinical characteristics

For the study, we collected the records of 2840 COVID-19 patients, 67 MERS patients, of which some may be the suspected cases, 43 SARS suspected patients, and 87 other HCoV patients. Table 1 shows the summary of clinical characteristics of the COVID-19, MERS, SARS, and HCoV patients. Among the COVID-19 patients, the mean age was 49.5 ± 25.9 and 1457 (51.3%) were males. Among the MERS patients, the mean age was 33.0 ± 21.7 and 35 (52.2%) were males whereas among the SARS and HCoV patients, the mean age of patients was 45.6 ± 30.9 and 10.6 ± 17.9, and 31 (72.1%) and 46 (52.9%) were males, respectively. From the comparison, COVID-19 group was significantly more likely to experience cancer (29.5% vs. 3.0%, p < 0.001) and gastrointestinal diseases (14.4% vs. 1.5%, p < 0.01) compared to the MERS group, whereas a higher proportion of SARS patients reported cerebrovascular (11.6% vs. 2.6%, p < 0.001), hepatic disease (11.6 % vs. 4.6 %, p < 0.05), renal disease (16.3% vs. 6.4%, p < 0.01) than the COVID-19 group. In symptom presentation, MERS and HCoV patients experienced fever (79.1% vs. 33.0%, p < 0.001 and 56.3% vs. 33.0%, p < 0.001, respectively) and upper respiratory infections (28.4% vs 0.8%, p < 0.001 and 34.5% vs 0.8%, p < 0.001) more frequently than the COVID-19 patients.

**Table 1.** Summary of clinical characteristics between the HCoVs, SARS, MERS, COVID-19 groups. Data are presented as the mean ± standard deviation for the continuous variables, otherwise the number of patients and percentage

| Characteristic (unit) | COVID-19 (n=2840) | MERS-CoV (n=67) | p-val | SARS-CoV-1 (n=43) | p-val | HCoV (n=87) | p-val |
| --- | --- | --- | --- | --- | --- | --- | --- |
| **Age (years)** |  |  |  |  |  |  |  |
| Total | 49.48 ± 25.94 | 33.04 ± 21.74 | <.0001 | 45.64 ± 30.93 | 0.422 | 10.59 ± 17.89 | <.0001 |
| 0-19 | 434 (15.28) | 19 (28.35) | 0.0035 | 13 (30.23) | 0.0072 | 72 (82.76) | <0.0001 |
| 20-39 | 563 (19.82) | 21 (31.34) | 0.020 | 3 (6.98) | 0.033 | 8 (9.20) | 0.014 |
| 40-59 | 561 (19.75) | 19 (28.35) | 0.082 | 8 (18.60) | 0.851 | 2 (2.30) | <0.0001 |
| 60-79 | 941 (33.13) | 7 (10.45) | <0.001 | 14 (32.56) | 0.937 | 5 (5.74) | <0.0001 |
| 80+ | 341 (12.00) | 1 (1.49) | 0.003 | 5 (11.63) | 0.939 | 0 (0.00) | NA |
| **Gender** |  |  |  |  |  |  |  |
| Male | 1457 (51.30) | 35 (52.23) | 0.879 | 31 (72.09) | 0.0068 | 46 (52.87) | 0.773 |
| **Comorbidities** |  |  |  |  |  |  |  |
| Total | 1376 (48.45) | 7 (10.45) | <0.0001 | 26 (60.47) | 0.118 | 31 (35.63) | 0.018 |
| Diabetes | 124 (4.36) | 1 (1.49) | 0.366 | 3 (6.98) | 0.436 | 1 (1.15) | 0.182 |
| Cardiovascular disease | 214 (7.53) | 0 (0.00) | NA | 6 (13.95) | 0.116 | 12 (13.79) | 0.031 |
| Cerebrovascular disease | 73 (2.57) | 0 (0.00) | NA | 5 (11.63) | 0.00028 | 2 (2.30) | 1.000 |
| Malignancy | 837 (29.47) | 2 (2.98) | <0.0001 | 8 (18.60) | 0.120 | 9 (10.34) | 0.00011 |
| Hepatic disease | 132 (4.65) | 0 (0.00) | NA | 5 (11.63) | 0.033 | 1 (1.15) | 0.185 |
| Gastrointestinal disease | 410 (14.44) | 1 (1.49) | 0.00061 | 9 (20.93) | 0.230 | 6 (6.90) | 0.047 |
| Renal disease | 183 (6.44) | 4 (5.97) | 1.000 | 7 (16.27) | 0.099 | 3 (3.45) | 0.259 |
| Thyroidal disease | 30 (1.06) | 1(1.49) | 0.516 | 1 (2.32) | 0.374 | 2 (2.30) | 0.246 |
| Musculoskeletal disease | 63 (2.22) | 0 (0.00) | NA | 1 (2.32) | 0.622 | 8 (9.20) | <0.0001 |
| **Symptoms** |  |  |  |  |  |  |  |
| Total | 1333 (46.94) | 63 (94.03) | <0.0001 | 17 (39.53) | 0.334 | 67 (77.01) | <0.0001 |
| Fever | 939 (33.06) | 53 (79.10) | <0.0001 | 9 (20.9) | 0.093 | 49 (56.32) | <0.0001 |
| Cough | 102 (3.59) | 8 (11.94) | 0.00040 | 3 (6.98) | 0.205 | 6 (6.90) | 0.107 |
| Dyspnea | 128 (4.51) | 2 (2.99) | 0.768 | 6 (13.95) | 0.0035 | 5 (5.75) | 0.584 |
| Chest pain | 63 (2.22) | 0 (0.00) | NA | 1 (2.32) | 0.622 | 0 (0.00) | NA |
| Sore throat | 51 (1.80) | 3 (4.48) | 0.127 | 0 (0.00) | NA | 1 (1.15) | 1.000 |
| Gastrointestinal symptoms | 193 (6.80) | 1 (1.49) | 0.129 | 2 (4.65) | 1.000 | 6 (6.90) | 0.971 |
| Upper respiratory infection | 23 (0.81) | 19 (28.36) | <0.0001 | 2 (4.65) | 0.052 | 30 (34.48) | <0.0001 |

#### Laboratory findings

Laboratory findings of each disease group are summarized in Table 2. Overall, COVID-19 patients showed increases in the level of creatinine and prothrombin time, and together with the SARS groups, experienced the elevated levels of alkaline phosphatase, total bilirubin, aminotransferases, and blood urea nitrogen. MERS group reported a high level of creatinine, whereas the HCoV group reported an increase in the level of alkaline phosphatase. All groups experienced high levels of fibrinogen. In complete blood cell counts, the levels of hemoglobin (13.18 ± 2.12 vs. 10.87 ± 2.47, p < 0.001), hematocrit (39.66 ± 5.59 vs. 33.03 ± 7.24, p < 0.001), and red blood cell count (4.54 ± 0.73 vs. 3.58 ± 0.85, p < 0.001) were significantly higher in the MERS group, whereas lymphocyte count (38.90 ± 27.00 vs. 18.44 ± 17.10, p < 0.001), red blood cell count (3.80 ± 0.75 vs. 3.58 ± 0.85, p < 0.05), eosinophil count (4.38 ± 6.09 vs. 1.37 ± 2.61, p < 0.01) were significantly higher in the HCoV group. Within the kidney function, blood urea nitrogen level was significantly higher in COVID-19 group than the MERS group and SARS group (23.38 ± 20.07 vs. 15.78 ± 13.80, p < 0.01 and 23.38 ± 20.07 vs. 12.90 ± 11.77, p < 0.001, respectively) and a significantly higher level of creatinine than the HCoV group (1.33 ± 1.77 vs. 0.58 ± 0.64, p < 0.01). eGFR was significantly higher in the MERS and SARS groups compared to the COVID-19 group (105.50 ± 61.10 vs. 78.27 ± 40.62, p < 0.001 and 107.50 ± 48.12 vs. 78.27 ± 40.62, p < 0.05, respectively).

**Table 2.** Summary of laboratory findings between the disease groups. Data are presented as the mean ± standard deviation for the continuous variables, otherwise the number of patients and percentage. Hb: Hemoglobain, Hct: Hematocrit, RBC: Red blood cell, WBC: White blood cell, PLT: Platelet, PCT: Procalcitonin, AST: Aspartate Aminotransferase, ALT: Alanine Aminotransferase, BUN: Blood urea nitrogen, eGFR: Estimated glomerular filtration rate, aPTT: Partial thromboplastin time, PT: Prothrombin time

| Characteristic (unit) | COVID-19 (n=2840) | MERS-CoV (n=67) | p-val | SARS-CoV-1 (n=43) | p-val | HCoV (n=87) | p-val |
| --- | --- | --- | --- | --- | --- | --- | --- |
| **Complete blood count** |  |  |  |  |  |  |  |
| Hb (g/dL) | 10.87 (2.47) | 13.18 (2.12) | <0.0001 | 10.81 (2.92) | 0.998 | 11.07 (2.19) | 0.911 |
| Hct (%) | 33.03 (7.24) | 39.66 (5.59) | <0.0001 | 31.50 (8.37) | 0.650 | 32.45 (6.56) | 0.846 |
| Lymphocyte (%) | 18.44 (17.1) | 18.93 (11.75) | 0.993 | 19.30 (15.10) | 0.984 | 38.90 (27.00) | <0.0001 |
| RBC ($10^6/\mu\text{L}$) | 3.58 (0.85) | 4.54 (0.73) | <0.0001 | 3.46 (0.99) | 0.872 | 3.80 (0.75) | 0.037 |
| WBC ($10^3/\mu\text{L}$) | 10.12 (15.21) | 8.10 (4.11) | 0.839 | 9.19 (6.94) | 0.984 | 5.64 (6.02) | 0.055 |
| Eosinophil (%) | 1.37 (2.61) | 1.20 (1.32) | 0.849 | 2.13 (4.71) | 0.761 | 4.38 (6.09) | 0.0020 |
| PCT (%) | 0.20 (0.11) | 0.20 (0.09) | 1.000 | 0.17 (0.09) | 0.142 | 0.19 (0.14) | 0.825 |
| PLT ($10^3/\mu\text{L}$) | 192.36 (120.45) | 217.00 (100.12) | 0.377 | 176.45 (114.49) | 0.819 | 178.95 (152.55) | 0.850 |
| **Liver function** |  |  |  |  |  |  |  |
| Albumin (g/dL) | 3.53 (0.74) | 4.17 (0.52) | <0.0001 | 2.94 (0.52) | <0.0001 | 3.76 (0.58) | 0.063 |
| Alkaline phosphatase (U/L) | 152.34 (191.25) | 89.98 (56.58) | 0.201 | 111.64 (76.22) | 0.460 | 132.41 (88.79) | 0.921 |
| Bilirubin, total (mg/dL) | 1.39 (2.35) | 0.67 (0.33) | <0.0001 | 2.37 (5.65) | 0.742 | 0.66 (0.46) | <0.0001 |
| AST (U/L) | 85.58 (440.14) | 33.50 (34.94) | 0.896 | 108.21 (261.10) | 0.993 | 38.40 (29.90) | 0.889 |
| ALT (U/L) | 54.25 (163.63) | 27.70 (24.84) | 0.772 | 92.82 (169.53) | 0.588 | 34.00 (31.26) | 0.841 |
| **Kidney function** |  |  |  |  |  |  |  |
| BUN (mg/dL) | 23.38 (20.07) | 15.80 (13.78) | 0.0044 | 28.50 (36.30) | 0.794 | 12.90 (11.77) | <0.0001 |
| Creatinine (mg/dL) | 1.33 (1.77) | 1.07 (1.12) | 0.801 | 1.03 (0.94) | 0.999 | 0.58 (0.64) | 0.0090 |
| eGFR | 78.27 (40.62) | 105.50 (61.10) | 0.0039 | 107.50 (48.12) | 0.023 | 70.79 (36.94) | 0.927 |
| **Coagulation function** |  |  |  |  |  |  |  |
| aPTT (sec) | 31.14 (12.68) | 31.85 (4.65) | 0.996 | 33.70 (7.58) | 0.883 | 31.18 (2.86) | 1.000 |
| Fibrinogen (mg/dL) | 400.28 (134.49) | 454.00 (122.64) | 0.391 | 458.81 (152.01) | 0.391 | 407.70 (132.59) | 0.999 |
| PT (sec) | 15.16 (10.50) | 13.00 (1.94) | 0.837 | 13.10 (2.69) | 0.881 | 12.74 (0.89) | 0.885 |

We performed an additional association analysis between the ABO blood type and coronavirus infections. As shown in Table 3, there were no statistically significant differences between the COVID-19, SARS, and HCoV groups by blood types. Due to the missing number of MERS patients who performed ABO blood test, we observed low p-values of less than 0.05 between the COVID-19 and MERS group comparison.

#### ABO blood type comparisons

**Table 3.** Summary of ABO blood type counts in each disease group

| Characteristic | COVID-19<br>(n=2840) | MERS-CoV<br>(n=67) | p-val | SARS-CoV-1<br>(n=43) | p-val | HCoV<br>(n=87) | p-val |
| --- | --- | --- | --- | --- | --- | --- | --- |
| **Blood type** |  |  |  |  |  |  |  |
| A | 596 (20.99) | 1 (1.49) | <0.0001 | 14 (32.56) | 0.065 | 18 (20.69) | 0.947 |
| B | 441 (15.53) | 2 (2.99) | 0.0028 | 10 (23.26) | 0.166 | 17 (19.54) | 0.310 |
| AB | 194 (6.83) | 0 (0.00) | NA | 2 (4.65) | 0.766 | 5 (5.75) | 0.692 |
| O | 470 (16.55) | 4 (5.97) | 0.018 | 7 (16.28) | 0.962 | 11 (12.6) | 0.333 |

### II. Comparative analyses of the COVID-19 severity

#### Demographic and clinical characteristics

After dividing the COVID-19 patients by the WHO disease severity criteria^18^, as demonstrated in Figure 1, 2596 people belonged to the mild group (91.4%) and 244 (8.6%) belonged to the non-mild group.

**Figure 1.**
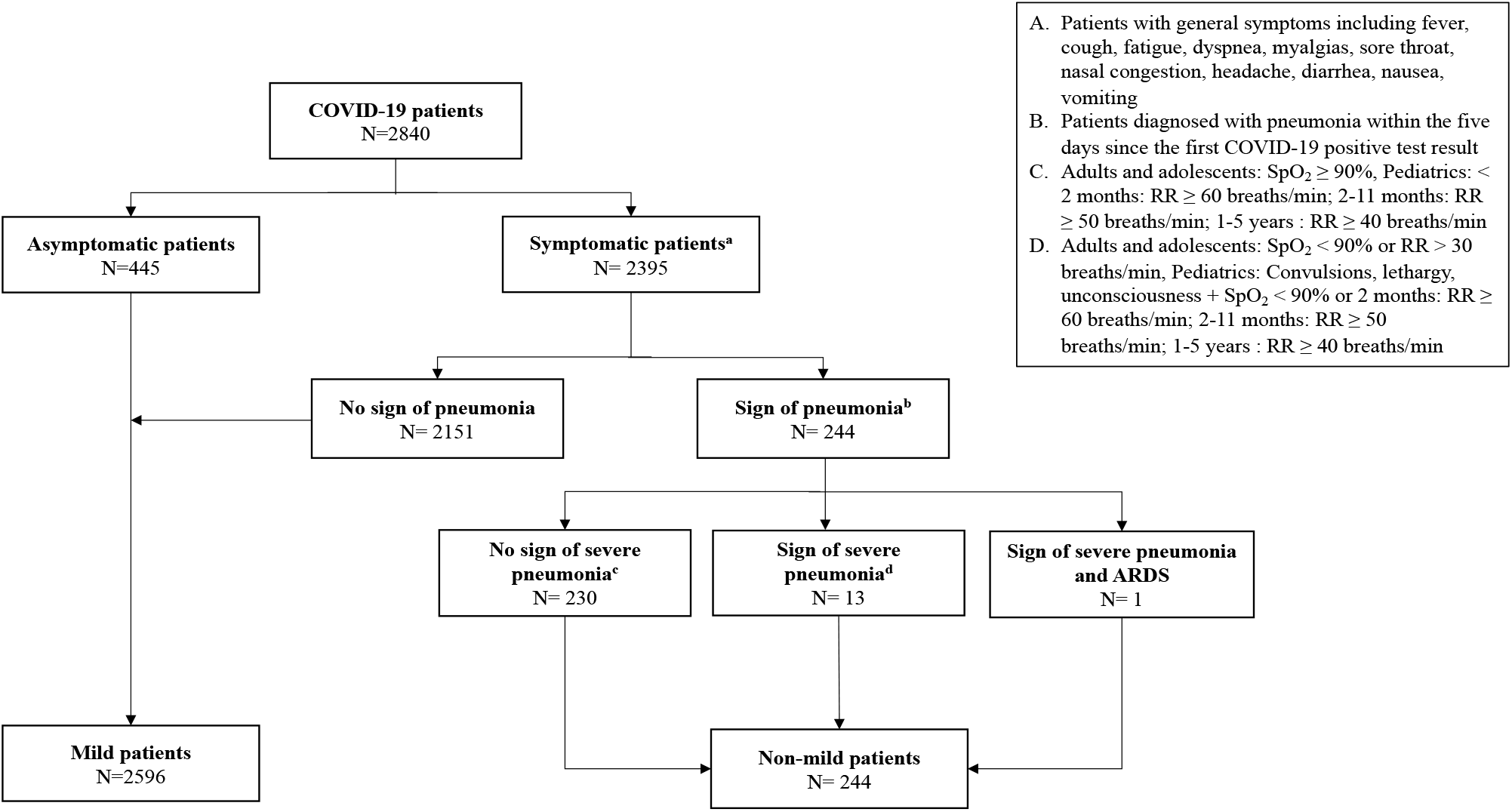
Flowchart of establishing cohorts by WHO guideline^18^

In Table 4, the mean age of mild group was 47.9 ± 25.7 and 1298 (50.0%) were males, and the mean age of non-mild group was 66.7 ± 22.6 and 159 (65.2%) were males. There were total 1376 COVID-19 patients with the comorbidities, of which 1240 and 136 patients belonged to the mild and non-mild group, respectively. By comparison, non-mild COVID-19 group presented significantly more comorbidities upon their hospital visits (55.7% vs. 47.8%, p < 0.05), including cardiovascular disease (13.1 % vs. 7.0%, p < 0.001), cerebrovascular disease (4.9% vs. 2.3%, p < 0.05), and renal disease (16.4% vs. 5.5 %, p < 0.01). For symptoms, non-mild group had significantly higher proportion of individuals reporting dyspnea (19.3% vs. 3.1%, p < 0.001), chest pain (5.7% vs. 1.9%, p < 0.001), and upper respiratory infections (2.5% vs. 0.7%, p < 0.001).

**Table 4.** Summary of clinical characteristics between the COVID-19 severity groups. Data are presented as the mean ± standard deviation for the continuous variables, otherwise the number of patients and percentage

| Characteristic (unit) | COVID-19<br>(n=2840) | Mild<br>(n=2596) | Non-mild<br>(n=244) | p-val |
| --- | --- | --- | --- | --- |
| **Gender** |  |  |  |  |
| Male | 1457 (51.30) | 1298 (50.00) | 159 (65.16) | <0.0001 |
| **Age (years)** |  |  |  |  |
| Total | 49.48 ± 25.94 | 47.86 ± 25.65 | 66.75 ± 22.55 | <0.0001 |
| 0-19 | 434 (15.28) | 415 (15.99) | 19 (7.79) | 0.00067 |
| 20-39 | 563 (19.82) | 555 (21.37) | 8 (3.28) | <0.0001 |
| 40-59 | 561 (19.75) | 536 (20.65) | 25 (10.25) | <0.0001 |
| 60-79 | 941 (33.13) | 828 (31.90) | 113 (46.31) | <0.001 |
| 80+ | 341 (12.00) | 262 (10.09) | 79 (32.38) | <0.0001 |
| **Comorbidities** |  |  |  |  |
| Total | 1376 (48.45) | 1240 (47.77) | 136 (55.74) | 0.017 |
| Diabetes | 124 (4.36) | 115 (4.43) | 9 (3.69) | 0.588 |
| Cardiovascular disease | 214 (7.53) | 182 (7.01) | 32 (13.11) | 0.00055 |
| Cerebrovascular disease | 73 (2.57) | 61 (2.35) | 12 (4.92) | 0.015 |
| Malignancy | 837 (29.47) | 758 (29.20) | 79 (32.38) | 0.298 |
| Hepatic disease | 132 (4.65) | 130 (5.07) | 2 (0.82) | 0.0012 |
| Gastrointestinal disease | 410 (14.44) | 390 (15.02) | 20 (8.20) | 0.004 |
| Renal disease | 183 (6.44) | 143 (5.51) | 40 (16.39) | <0.0001 |
| Thyroidal disease | 30 (1.06) | 30 (1.16) | 0 (0.0) | NA |
| Musculoskeletal disease | 63 (2.22) | 55 (2.12) | 8 (3.28) | 0.239 |
| **Symptoms** |  |  |  |  |
| Total | 1333 (46.94) | 1234 (47.53) | 99 (40.57) | 0.037 |
| Fever | 939 (33.06) | 895 (34.48) | 44 (18.03) | <0.0001 |
| Cough | 102 (3.59) | 97 (3.74) | 5 (2.05) | 0.176 |
| Dyspnea | 128 (4.51) | 81 (3.12) | 47 (19.26) | <0.0001 |
| Chest pain | 63 (2.22) | 49 (1.89) | 14 (5.74) | <0.0001 |
| Sore throat | 51 (1.80) | 51 (1.96) | 0 (0.0) | NA |
| Gastrointestinal symptoms | 193 (6.80) | 185 (7.13) | 8 (3.28) | 0.022 |
| Upper respiratory infection | 23 (0.81) | 17 (0.65) | 6 (2.46) | 0.0026 |

When comparing the mild and non-mild groups of COVID-19 patients, Table 5 shows a difference between the severity by the blood type A, with a significantly higher proportion of non-mild COVID-19 patients in type A compared to the mild group (30.7 % vs. 20.1%, p < 0.001).

#### ABO blood type comparisons

**Table 5.** Summary of ABO blood type counts in each severity group

| Characteristic | COVID-19<br>(n=2840) | Mild<br>(n=2596) | Non-mild<br>(n=244) | p-val |
| --- | --- | --- | --- | --- |
| **Blood type** |  |  |  |  |
| A | 596 (20.99) | 521 (20.07) | 75 (30.74) | <0.0001 |
| B | 441 (15.53) | 409 (15.76) | 32 (13.11) | 0.276 |
| AB | 194 (6.83) | 172 (6.63) | 22 (9.02) | 0.157 |
| O | 470 (16.55) | 428 (16.49) | 42 (17.21) | 0.770 |

#### Laboratory findings

Laboratory findings of each severity group are summarized in Table 6. From the table, we observed an increase in the count of white blood cell and the levels of blood nitrogen urea and fibrinogen within the non-mild group and an increase in the level of alkaline phosphatase within the mild group. Both severity groups reported elevated levels of aminotransferases and creatinine. Compared to the mild group, non-mild group showed statistically higher levels of white blood cell count (11.13 ± 9.44 vs. 9.99 ± 0.16, p < 0.05), eosinophil count (1.93 ± 4.30 vs. 1.28 ± 2.25, p < 0.01), procalcitonin (0.21 ± 0.11 vs. 0.20 ± 0.11, p < 0.01), and platelet (208.26 ± 120.78 vs. 190.18 ± 120.26, p < 0.05). While in liver function measurements, the mild group showed higher levels of albumin (3.56 ± 0.74 vs. 3.30 ± 0.64, p < 0.001), alkaline phosphatase (156.52 ± 200.38 vs. 123.94 ± 106.64, p < 0.001), and total bilirubin (1.45 ± 2.47 vs. 0.94 ± 1.16, p < 0.001), they had lower levels of blood nitrogen urea (22.50 ± 19.56 vs. 29.56 ± 22.45, p < 0.001), creatinine (1.28 ± 1.71 vs. 1.78 ± 2.18, p < 0.001), and fibrinogen (392.81 ± 135.42 vs. 449.46 ± 117.01, p < 0.001) compared to the non-mild group.

**Table 6.** Summary of laboratory findings between the COVID severity groups. Data are presented as the mean ± standard deviation for the continuous variables, otherwise the number of patients and percentage. Hb: Hemoglobin, Hct: Hematocrit, RBC: Red blood cell, WBC: White blood cell, PLT: Platelet, PCT: Procalcitonin, AST: Aspartate Aminotransferase, ALT: Alanine Aminotransferase, BUN: Blood urea nitrogen, eGFR: Estimated glomerular filtration rate, aPTT: Partial thromboplastin time, PT: Prothrombin time

| Characteristic (unit) | Total | Mild | Non-mild | p-val |
| --- | --- | --- | --- | --- |
| **Complete blood count** |  |  |  |  |
| Hb (g/dL) | 10.97 (2.47) | 10.91 (2.47) | 10.58 (2.40) | 0.013 |
| Hct (%) | 33.03 (7.24) | 33.10 (7.25) | 32.58 (7.19) | 0.181 |
| Lymphocyte (%) | 18.44 (17.1) | 18.63 (0.17) | 17.16(16.44) | 0.109 |
| RBC ($10^6/\mu\text{L}$) | 3.58 (0.85) | 3.59 (0.85) | 3.47 (0.82) | 0.0061 |
| WBC ($10^3/\mu\text{L}$) | 10.12 (15.21) | 9.99 (0.16) | 11.13(9.44) | 0.043 |
| Eosinophil (%) | 1.37 (2.61) | 1.28 (2.25) | 1.93 (4.30) | 0.0090 |
| PCT (%) | 0.20 (0.11) | 0.20 (0.11) | 0.21 (0.11) | 0.0027 |
| PLT ($10^3/\mu\text{L}$) | 192.36 (120.45) | 190.18 (120.26) | 208.26 (120.78) | 0.0057 |
| **Liver function** |  |  |  |  |
| Albumin (g/dL) | 3.53 (0.74) | 3.56 (0.74) | 3.30 (0.64) | <0.0001 |
| Alkaline phosphatase (U/L) | 152.34 (191.25) | 156.52 (200.38) | 123.94 (106.64) | <0.0001 |
| Bilirubin, total (mg/dL) | 1.39 (2.35) | 1.45 (2.47) | 0.94(1.16) | <0.0001 |
| AST (U/L) | 85.58 (440.14) | 88.08(456.98) | 67.99 (295.15) | 0.274 |
| ALT (U/L) | 54.25 (163.63) | 53.57(143.78) | 59.05 (2644.81) | 0.705 |
| **Kidney function** |  |  |  |  |
| BUN (mg/dL) | 23.38 (20.07) | 22.50 (19.56) | 29.56 (22.45) | <0.0001 |
| Creatinine (mg/dL) | 1.33 (1.77) | 1.28(1.71) | 1.78 (2.18) | <0.0001 |
| eGFR | 78.27 (40.62) | 79.41(39.98) | 70.66 (43.97) | 0.00041 |
| **Coagulation function** |  |  |  |  |
| aPTT (sec) | 31.14 (12.68) | 31.12(12.46) | 31.22 (14.06) | 0.909 |
| Fibrinogen (mg/dL) | 400.28 (134.49) | 392.81(135.42) | 449.46 (117.01) | <0.0001 |
| PT (sec) | 15.16 (10.50) | 15.15(11.11) | 15.25 (4.79) | 0.801 |

### III. Logistic regression

In order to observe the extent of common clinical characteristics among disease groups that may uniquely affect the prognosis of COVID-19 patients, we conducted a nominal logistic regression with the variables that showed the p values higher than 0.05 in two thirds of the comparative groups between the diseases and less than 0.05 in the analyses between the disease severity groups. We found no collinearity between the variables. Table 7 below shows the results from regression that has been adjusted for both age and gender. The univariate logistic regression showed that the male patients (OR: 1.66; CI: 1.29-2.13, p < 0.001) with ABO blood type A (OR: 1.80; CI: 1.40-2.31, p < 0.001), renal disease (OR: 3.27; CI: 2.34-4.55, p < 0.001), decreased creatinine (OR: 2.05; CI: 1.45-2.88, p < 0.001), and increased level of fibrinogen (OR: 1.59, CI: 1.21-2.09, p < 0.001) are associated with the higher risks of progressing towards the more severe stage whereas the patients with gastrointestinal symptoms (OR: 0.42; CI: 0.23-0.72, p < 0.01) and increased alkaline phosphatase (OR: 0.73; CI: 0.56-0.94, p < 0.05) are less likely to experience the severe prognosis of the disease. Age was not associated with risk of high severity.

**Table 7.** Nominal logistic regression adjusted for age and gender. Hb: Hemoglobin, WBC: White blood cell, PCT: Procalcitonin, PLT: Platelet. Normal ranges served as the reference values for all measurement variables

| Characteristic (unit) | OR (95% CI) | p-val |
| --- | --- | --- |
| **Age (years)** | 1.04 (1.02, 1.06) | 0.00087 |
| 0-19 | 2.74 (0.46, 16.04) | 0.265 |
| 20-39 | 1.18 (0.30, 4.53) | 0.808 |
| 40-59 | 0.82 (0.36, 1.85) | 0.636 |
| 60-79 | 0.80 (0.52, 1.23) | 0.311 |
| **Male, Sex** | 1.66 (1.29, 2.13) | <0.0001 |
| **Blood type (A)** | 1.80 (1.40, 2.31) | <0.0001 |
| **Dyspnea** | 5.36 (3.88, 7.41) | <0.0001 |
| **Renal disease** | 3.27 (2.34, 4.55) | <0.0001 |
| **Gastrointestinal symptoms** | 0.42 (0.23, 0.72) | 0.0032 |
| **Hb (g/dL)** |  |  |
| High | 2.17 (0.94, 4.70) | 0.057 |
| Low | 0.98 (0.73, 1.32) | 0.882 |
| **WBC ($10^3/\mu\text{L}$)** |  |  |
| High | 1.38 (1.06, 1.80) | 0.016 |
| Low | 1.72 (1.18, 2.49) | 0.0044 |
| **Eosinophil (%)** |  |  |
| High | 1.50 (0.76, 2.80) | 0.217 |
| Low | 0.94 (0.74, 1.20) | 0.628 |

| PCT (%) |  |  |
| --- | --- | --- |
| High | 0.88 (0.40, 1.98) | 0.753 |
| Low | 0.73 (0.54, 0.99) | 0.045 |
| PLT (10<sup>3</sup>/μL) |  |  |
| High | 2.25 (1.05, 4.60) | 0.030 |
| Low | 0.97 (0.68, 1.37) | 0.845 |
| Alkaline Phosphatase (U/L) |  |  |
| High | 0.73 (0.56, 0.94) | 0.015 |
| Low | 0.36 (0.06, 1.30) | 0.182 |
| Creatinine (mg/dL) |  |  |
| High | 0.92 (0.68, 1.23) | 0.575 |
| Low | 2.05 (1.45, 2.88) | <0.0001 |
| Fibrinogen (mg/dL) |  |  |
| High | 1.59 (1.21, 2.09) | 0.00084 |
| Low | 0.81 (0.36, 1.63) | 0.574 |

Figure 2 shows the proportion of patients with risk factors in non-mild group, of which elevated fibrinogen level (72.5%) and male sex (58.2%) most commonly appeared. When comparing the identical parameters in different disease groups, Figure 3 shows that the proportions of patients in male sex (72.1%), blood type A (32.6%), dyspnea, and renal disease were the highest in SARS group whereas the number of patients with elevated WBC was the highest in MERS group. The rates of elevated fibrinogen were similar in SARS and MERS groups with 25% and 26.2%, respectively.

**Figure 2.**
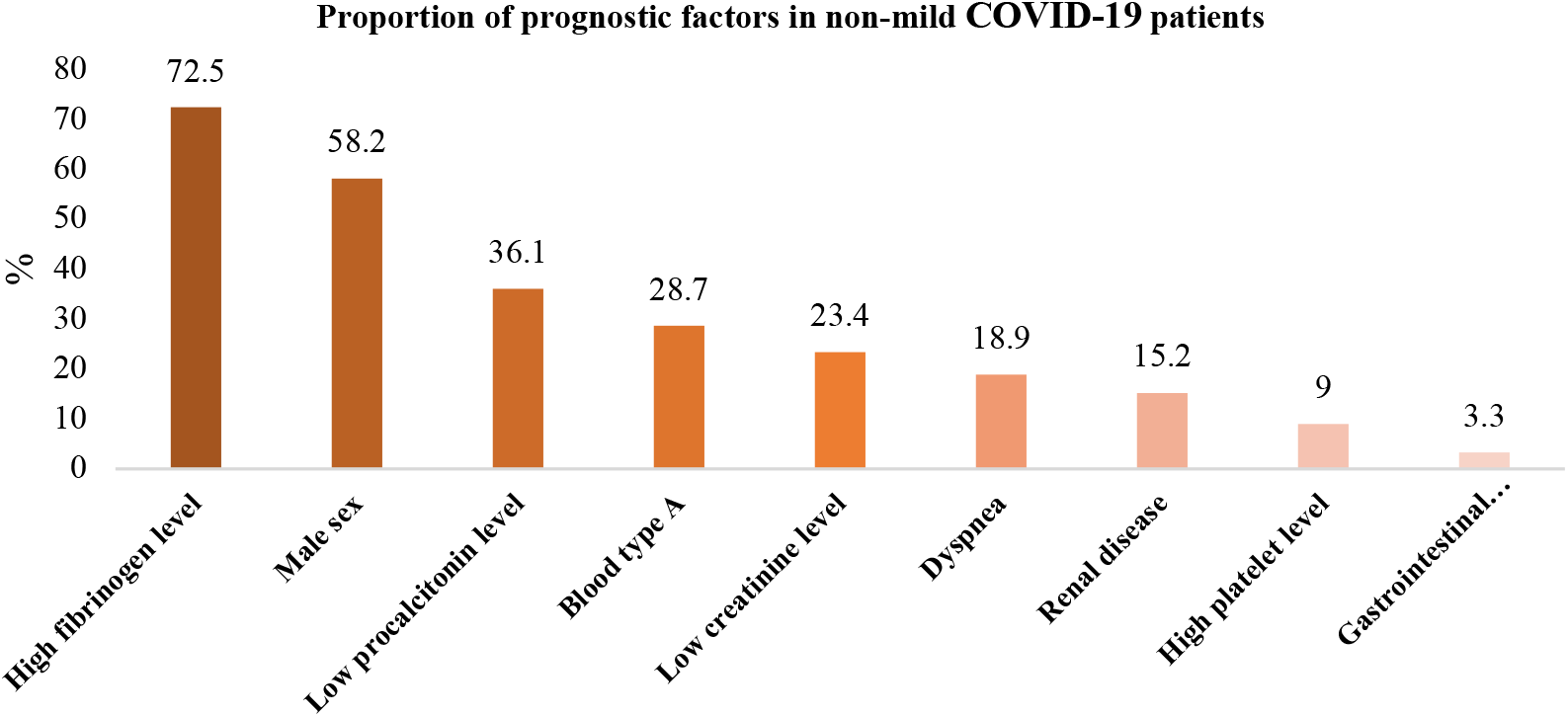
Proportion of prognostic factors in non-mild COVID-19 patients

**Figure 3.**
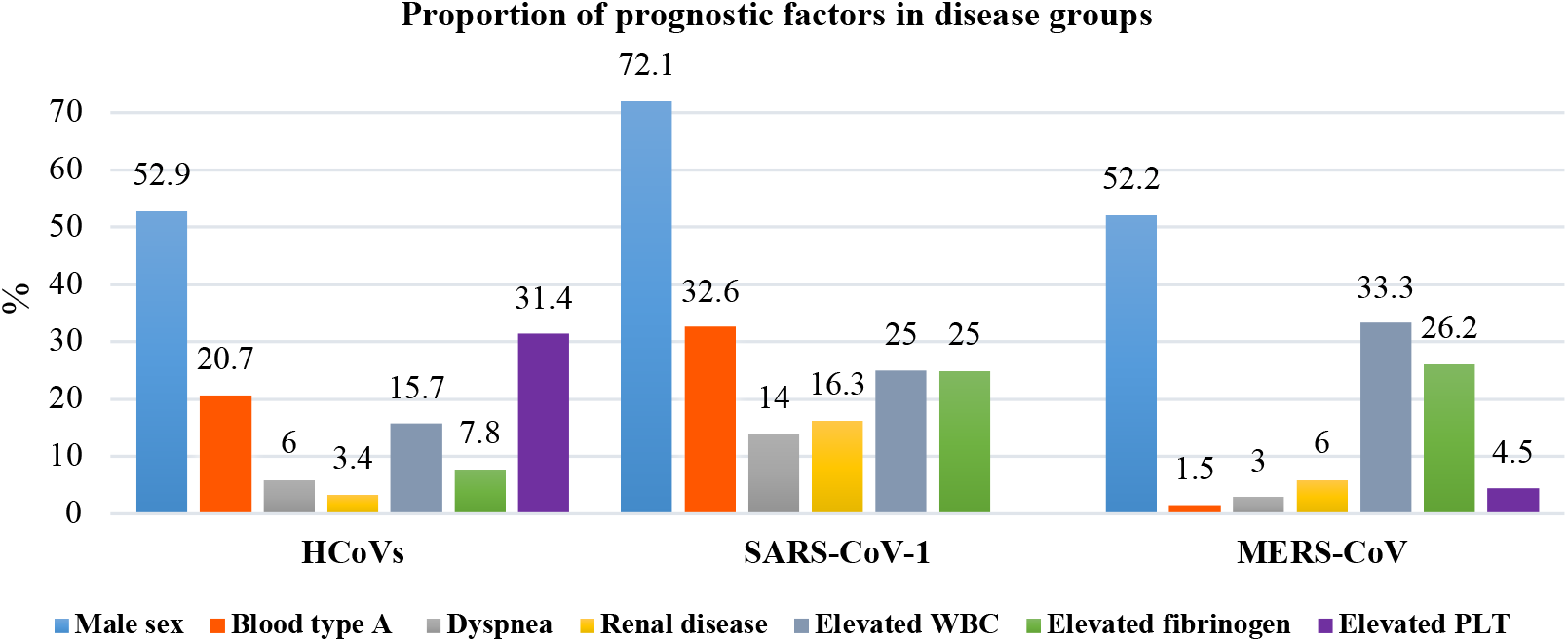
Proportion of prognostic factors in disease groups

## Discussion

The present retrospective cohort study examined the clinical resemblance between the coronavirus infected patients within South Korea and identified the common characteristics and risk factors that can potentially influence a severity of the novel COVID-19. In our findings, we discovered the most similarities in clinical features between the COVID-19 group, especially the non-mild COVID-19 patients, and SARS group, showing statistical similarities that range from 75-89.5% in their manifestations. Our observation between the SARS-CoV-2 and SARS-CoV-1, analogous to a previous study by Petrosillo et al^28^, maybe attributed to a resemblance of more than 80% in genomic sequences^29^. It is also to note that the two viruses utilize an angiotensin-converting enzyme 2 (ACE2) receptor, suggesting the common cellular binding mechanism as the basis of parallel pathogeneses in COVID-19 and SARS patients^30-32^.

Another important finding was that the male sex, blood type A, renal disease, and elevated level of fibrinogen were major risk factors in virus infections and severe prognosis of COVID-19. In line with our observation in the patient demographics, of which more than 50% of individuals were males, previous research on SARS data in Hong Kong showed enhanced fatality rate in men by 21.9% compared to women with 13.2%^33-34^ whereas the South Korean and Saudi Arabic MERS-CoV data predominantly reflected male sex in infected population, with the respective frequencies of 65.2% and 59.1%^35^. One theory for a higher risk in males includes greater circulating ACE2 levels than in women, which creates a sex-dependent difference in virus infection^36-39^, evidenced by a positive correlation of susceptibility of male mice to SARS-CoV and virus titers observed in a vitro study^39^. However, sex-dependent discrepancy is multifactorial, which would also require further investigations in relation to socioeconomic and behavioral elements.

In our study, we did not see a significant difference within a distribution of ABO blood types between the disease groups, yet the proportion of blood type A individual was significantly higher in the non-mild COVID-19 group. Such finding parallels to the meta-analysis on Spanish and Italian patient data where the odds for having severe COVID-19 were higher in A/AB groups as compared with B/O group^40-41^. While the research regarding the association between ABO blood type and coronavirus infection is relatively new, one potential mechanism of type A group’s susceptibility can be attributed to a strong affinity of SARS-CoV-2 spike protein to the carbohydrates, such as galactose in the A antigen, unlike a galactose amine that is present as end group saccharide in the B and O antigens^42-43^. Another speculation pertaining to severity risk difference between the type A and non-A patients involves a protective role of neutralizing anti-A antibody blocking the interaction of ACE2 and S protein^44^.

The kidney failures were evident in all disease groups and were shown to be the variable with the second-highest risk rate next to dyspnea. Firstly, ACE2, which is present in multiple organs has also been found to be highly expressed on renal cells, facilitating SARS-CoV-2 entry and consequent acute tubular injury, and occasionally collapsing focal segmental glomerulopathy in the kidney tissue^45-47^. Studies suggest a decrease in the ACE2 as a result of viral attachment to the receptor and the consequent elevation in the level of AngII and AT1R l stimulation, which induce an aggravation of respiratory distress in COVID-19 patients^48-52^. Further, CKD patients are predisposed to the poorer outcomes—their immunosuppressed state and chronic systemic inflammation can increase the risks of developing more severe complications and mortality^53-54^. Likewise, in our data, chronic kidney disease (CKD) and acute kidney injury (AKI) were the two most frequently reported diagnoses.

Of the major laboratory findings, we noticed the overall increase fibrinogen in our cohorts, especially within the non-mild COVID-19 group. Elevated fibrinogen level can be an indicator for hypercoagulability due to viral pneumonia, which is a common complication in coronavirus infections^55-57^. A literature review by Abou-Ismail et al revealed the prothrombotic traits in SARS-CoV-2 patients globally^58^, which is also evidenced by our records that showed occurrences of thrombotic manifestation including pulmonary embolism, myocardial infarction, ischemic stroke, and deep vein thrombosis (DVT) in COVID-19 and SARS patients. Coagulopathy in coronavirus infected patients has been linked to a heightened inflammatory response that typically leads to thrombo-inflammation. The inflammation is suspected to begin in lungs damaging the pulmonary vasculature that leads to thrombosis in the early stage of the disease^58-66^.

To conclude, our study detected the highest resemblance between the COVID-19 and SARS groups. Further, clinical manifestations that were present in SARS group were linked to the severity of COVID-19. The regression model, upon applying the common variables between the disease groups, indicated male sex, blood type A, dyspnea, renal disease, and elevated levels of white blood cell, platelet, fibrinogen and alkaline phosphatase, and decreased creatinine level as the risk factors of severe COVID-19. Our study had several limitations; first, there existed missing data, which may explain a disparity in our creatinine analyses. Considering the relative nature of logistic regression, we speculate the model to presume the contradictory correlations between the disease severity and creatinine level—only 16% of the individuals with high creatinine level belonged to the non-mild group, yet it should be noted that the number of admitted severe patients in Seoul National University Hospital was relatively low. Further, there were missing PCR data available on our database— although we utilized the diagnoses that existed on the system, there may involve the suspected cases, especially within the SARS group. Therefore, this study provides the limited clinical observations that represent the disease groups. Second, the study population was limited to the patients at a single national hospital due to the limited study period. Thus, the results in this study should not be generalized fully to other populations and ought to be considered with a caution.

Yet, we maximized the benefit of utilizing the anonymized EMR data integrated into CDM, which now is expanding in its scope globally for the vitalization of medical research^67^. Upon choosing a database, we ensured that the domains from the CDW were equally available in our CDM and that there were no gaps between the amount of data in each database with the frequent extract, transform, and load (ETL) process. Thus, through the use of CDM, we were able to reflect the real-world medical challenges in managing coronavirus infection, providing the most updated clinical dynamics. To our knowledge, this is the first study that attempts to clinically characterize and compare four major human coronavirus infections as well as to identify the common risk factors found in previous viral infections that may influence the prognosis of novel COVID-19. With our analyses that applied the variables of both the statistical and clinical significance, we expect the results from the present study to provide clinical insights that can be used as a basis for predicting the prospective clinical representations by yet another coronavirus-derived illness in near future and to serve the guidelines for clinical treatments of current patients. However, there still left a great uncertainty around the set of symptoms and complications from coronavirus, and it is absolutely crucial to further investigate for attaining a better understanding of the disease. For future study, therefore, we recommend performing a cross-country clinical characterization and survival analysis to determine risk factors that are particular to ethnic group, which shall aid in developing more specific control and prevention schemes for each country. Moreover, it will be insightful to analyze the manifestations at a multidimensional perspective, including the patients’ socioeconomic background, nutritional status, and behavioral habit that might exert an influence on the disease outcomes, with the integration of additional data from multiple institutions.

## Data Availability

The data that support the findings of this study are available on request from the corresponding author, Kwangsoo Kim. The data are not publicly available as they contain information that could compromise the privacy of research participants.

## Acknowledgement

This work was supported by the Technology Innovation Program (20003883, Technology development on CDM-based biohealth integrated data network extension) funded by the Ministry of Trade, Industry & Energy (MOTIE, Korea) and a grant of the Korea Health Technology R&D Project through the Korea Health Industry Development Institute (KHIDI), funded by the Ministry of Health & Welfare, Republic of Korea (grant number: HI19C1234). The funders of the study had no role in study design, data collection, data analysis, data interpretation, or writing of the report.

## Appendix 1. Normal range

**Table 1.**
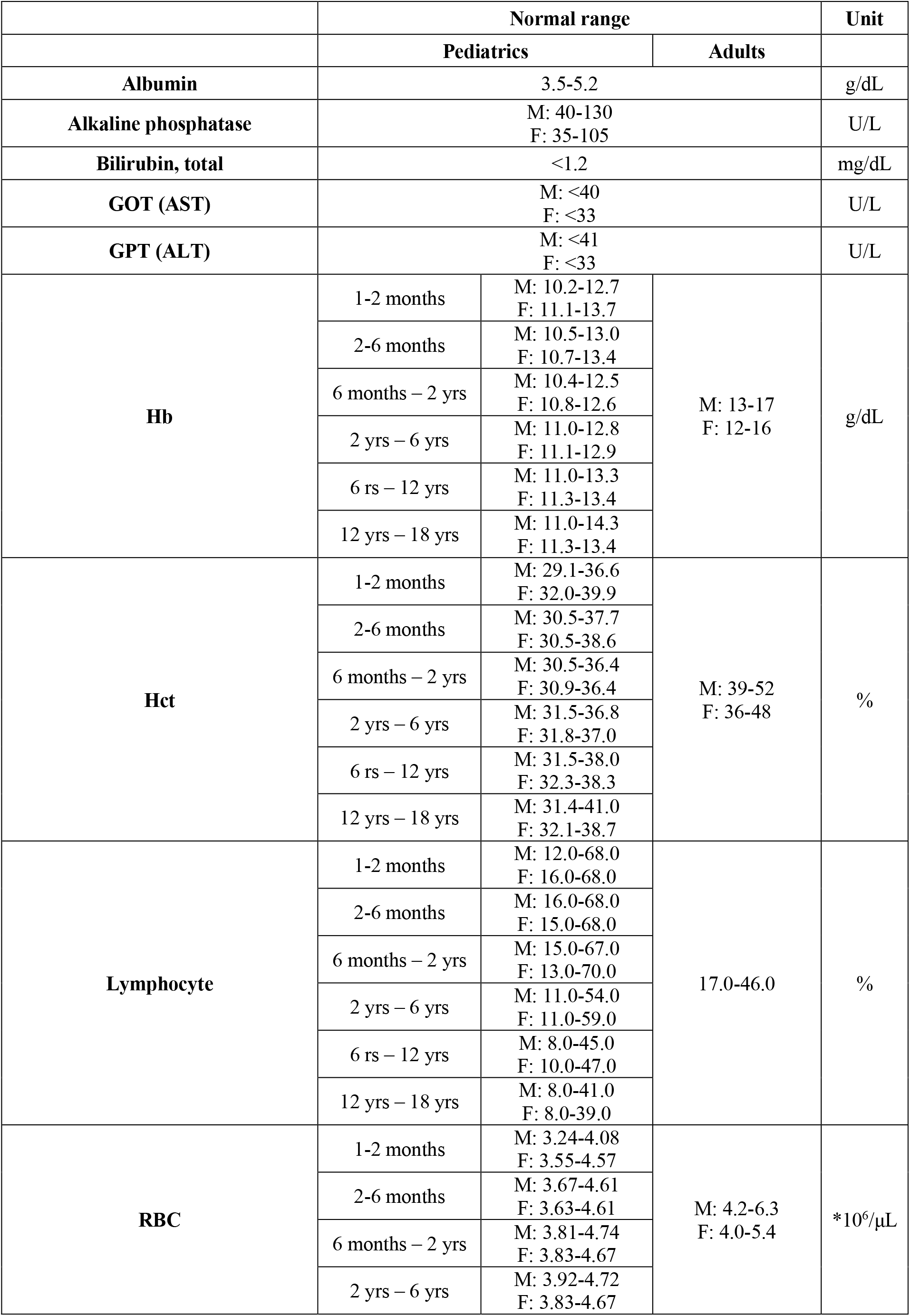
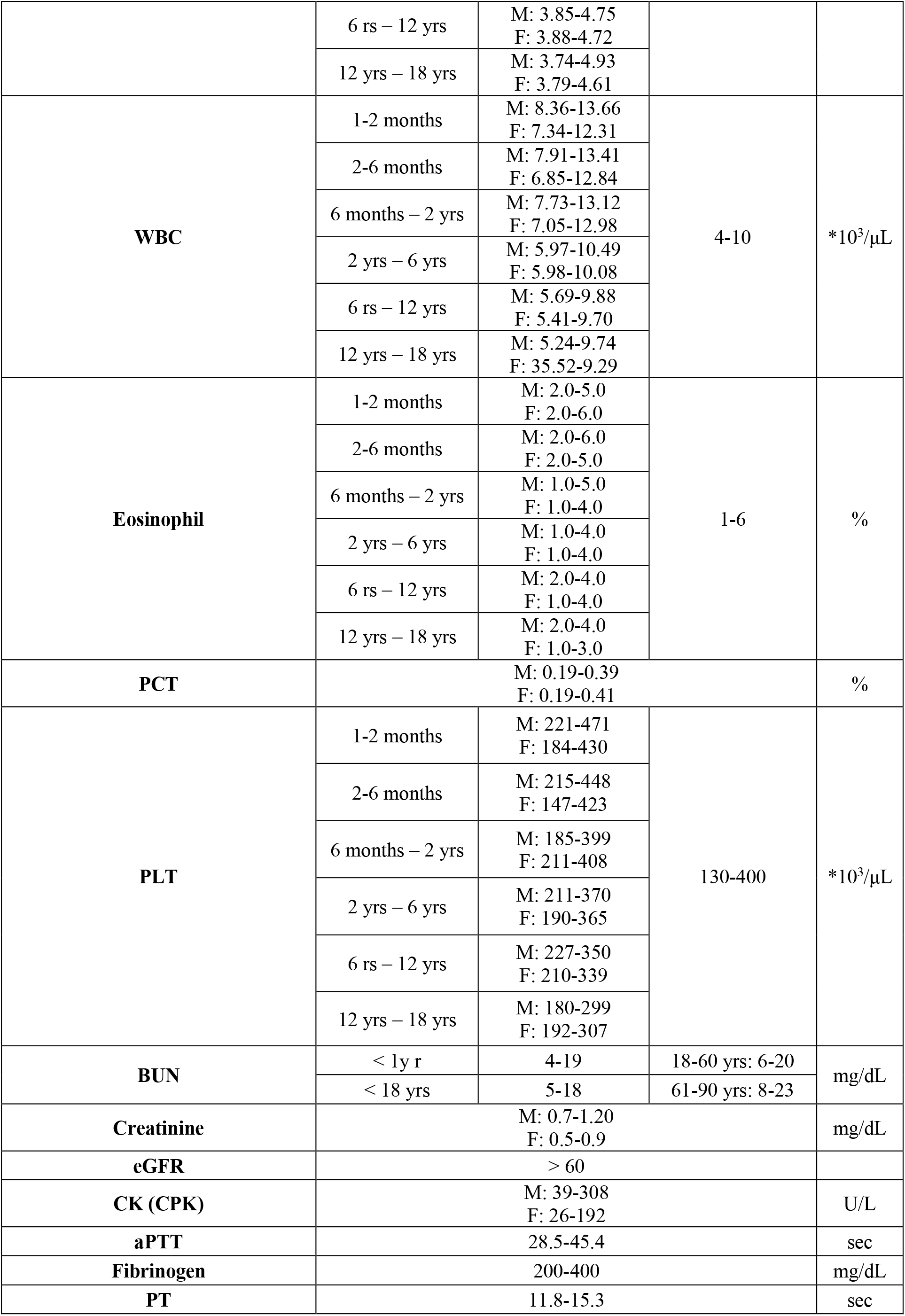
Normal ranges used for the continuous variables^19^.

